# Spatial variation in the non-use of modern contraception and its predictors in Bangladesh

**DOI:** 10.1101/2023.03.23.23287644

**Authors:** Md Nuruzzaman Khan, Melissa L. Harris

## Abstract

**Objective:** We assessed spatial variations in the non-use of modern contraception in Bangladesh, and identified associated individual, household and community level factors.

**Methods:** We analysed data from 16,135 women extracted from the 2017/18 Bangladesh Demographic and Health Survey. The study outcome was non-use of modern contraception (yes, no), and the explanatory variables were factors at the individual (e.g., women’s education), household (e.g., husband education) and community level (e.g., community level poverty and illiteracy). Moran’s I statistics was applied to examine whether any geographical heterogeneity in non-use of modern contraception exists in Bangladesh. The Gettis-ord Gi^*^ was calculated to measure how spatial autocorrelation differed across study locations. A geographically weighted regression model was used to assess the relationship of non-use of modern contraception at the cluster level.

**Results:** Overall, 42.8% (95% CI, 41.6-43.8) of respondents reported non-use of modern contraception in Bangladesh with a significant variation across geographical locations (p<0.001). Clusters of high non-use of modern contraception (hot spots) were found mostly located in the Sylhet, Barishal and part of the Chattogram divisions while clusters of low use of modern contraception(cold spots) were mostly located in the Rangpur, Mymensingh and part of the Rajshahi divisions. The likelihood of non-use of modern contraception was strongest among women and parteners with low levels of education. Other risk factors analysed including partner’s occupation, community-level illiteracy and poverty had varying effects on the non-use of modern contraception across the locations (clusters) of the country included in the survey.

**Conclusion:** Prevalence of, and risk factors for, modern contraception non-use in Bangladesh differed depending on geographical location. This suggests a need for targeted area-specific policies and programs to improve knowledge and uptake of modern contraception.

## Introduction

Three key targets of the Sustainable Development Goals (SDGs) focus on ensuring universal access to sexual and reproductive healthcare services (target 3.7) and reducing maternal (target 3.1) and child mortality (target 3.2) by 2030. Use of modern contraception is critical to achieving these targets [1]. However, nearly 55% of the world’s reproductive aged women (270 million) do not use modern contraception. A large proportion of these women live in Sub-Saharan Africa (75%) and Central (72%) and South Asia (58%) [2, 3]. Consequently, over 82% of the total 121 million global occurrence of unintended pregnancies occur in Asian and African regions [4]. In particular, around 2 million pregnancies occur in Bangladesh where around half of pregnancies are reported as unintended [5]. Given the prevalence of non-use of modern contraception in Bangaldesh is high (58%), this places women and children at significant risk of adverse maternal and child health consequences due to abortion and unplanned birth [6, 7]. Abortion due to unintended pregnancy has been found to be responsible for up to 13% of maternal deaths each year in LMICs [8, 9]. The rate is suspected to be higher for countries such as Bangladesh, where abortion is completely prohibited unless provided to save a woman’s life. Specific estimates however are scarce. Moreover, approximately 7 million women in LMICs are admitted to the hospital due to post-abortion complications including haemorrhage, sepsis, and injury that costs the economy US$ 553 million a year [8]. Maternal mortality in Bangladesh is also higher among women who choose to continue with their unintended pregnancies, largely due to lower use of intrapartum, birthing and postpartum care [7, 10-13]. Low use of maternal healthcare services is often compounded by individual level factors (e.g., depression, smoking) which increase the risk of adverse child health outcomes, including stillbirth, preterm birth, and low birth weight [7]. These are ongoing challenges in LMICs such as Bangladesh [7] and point to the need to address this disparity [1].

Previous studies from LMICs countries including Bangladesh have estimated the prevalence of modern contraception use, unmet need for contraception, unmet need for modern contraception, and the factors associated with these outcomes [14–20]. Although these studies have provided pertinent information that has been used to inform family planning policies, they have mostly provided broad population level evidence with significant difference of contraception use across areas but have failed to understand the nuances associated with geographical location [18]. Consequently, family planning policies and programs in LMICs, including Bangladesh, are mostly identical for the entire country with no area-specific programs available that consider the area-level factors that influence non-use of modern contraception [21, 22]. This poses difficulties in not only access to, but also uptake of modern contraception and often results in the misuse of manpower and resources. Consequently, this creates a further burden on the healthcare sector and prevents the ability to achieve potential success in the maternal and child health outcomes. These challenges cannot be overcome without identifying the areas where the non-use of contraception is highest and the associated risks factors.

To address these limitations and to assist policymakers in developing targeted evidence-based area-specific policies, we aimed to determine the spatial variations of non-use of modern contraception in Bangaldesh and identify associated individual-, households-, and community level factors.

## Methods

### Survey background and design

Data from the 2017/2018 Bangladesh Demographic and Health Survey (BDHS) were analysed. The BDHS is a nationally representative cross-sectional survey conducted as part of the Demographic and Health Survey Program. In Bangladesh, the BDHS is implemented by the National Institute of Population Research and Training and supervision of the survey’s administration and monitoring is carried out by the Ministry of Health and Family Welfare. The focus of this survey was on reproductive-aged married women who resided permanently in the selected households or stayed at the selected households on the night before the survey. The households were selected in two stages. At the first stage, a total of 675 area clusters were selected randomly from a list of 293,579 area clusters which was generated by the Bangladesh Bureau of Statistics as part of the 2011 Bangladesh National Population Census. A total of 672 area clusters were retained after excluding three area clusters due to flood. A total of 20,160 households were selected. and 19,457 households completed the surveys (96% response rate). In these selected households, there were 20,376 women, of which, 20,127 were interviewed (98.8% response rate). A detailed description of the sampling procedure is available elsewhere [5].

The 2017/18 BDHS also collected geospatial data, which included the latitudinal and longitudinal coordinates of each cluster included in the survey using a geographical pointing system. These data were linked to the survey data for analysis.

### Participants

For inclusion in this analysis, women had to be (i) not pregnant at the time of data collection, (ii) not in the lactational amenorrhea period and (iii) not planning to having children within two years of the survey. Women also had to have geospatial data available for linkage.

### Study variable

The study outcome variable was non-use of modern contraception. During the survey, eligible women were asked *“Are you currently doing something or using any method to delay or avoid getting pregnant?”* Women who responded “Yes” to this item were then asked, *“Which method are you using?”* and could select their method from the following list: the pill, injectable, implant, intrauterine device (IUD), condoms, female sterilization, male sterilization, periodic abstinence and withdrawal. A free text option was also provided to enable reporting the name of contraception if not included in the list. If a women reported multiple methods, the method she uses most frequently was selected. Responses were coded as “0” for use of modern contraception (i.e., the injectable, pill, implant, IUD, female sterilization, male sterilization, and condoms) and “1” for non-use of modern contraception (responded “no” to the first question) or for use of traditional methods (i.e., periodic abstinence or withdrawal) from the second question.

### Explanatory variables

The explanatory variables considered were factors at the individual-, household-, and community-level selected based on relevant published literature in LMICs including Bangladesh [14–20]. Individual level factors included were women’s age (≤19 years, 20-34 years, ≥35 years), education (illiterate, primary, secondary, higher) and working status (yes, no). Household level factors included were husband’s education (illiterate, primary, secondary, higher), husband’s occupation (agriculture, physical worker, services or business), number of children ever born (≤2 children, ≥3 children), types of family where women resided (nuclear, joint), preceding birth interval (≤ 2 years, 2–4 years, >4 years) and household wealth quintile (poorest, poor, middle, rich, richest). The factors at the community level included were place of residence (urban, rural), division (Barishal, Chattogram, Dhaka, Khulna, Mymensingh, Rajshahi, Rangpur and Sylhet), community-level illiteracy (low (≤ 20%), moderate (21.0-49.0) and high (≥ 50), community-level poverty (low (≤ 20%), moderate (21.0-49.0) and high (≥ 50), middle to richest community) and community level fertility (low(≤ 2.10), high (>2.10)). We calculated community level illiteracy, community level poverty, community level fertility variables using aggregated responses to the education, poorest and poorer wealth quintile and number of children ever born questions at the cluster level, respectively. The detailed calculation procedure has been published elsewhere [17].

### Statistical analysis

We first calculated the proportion of non-use of modern contraception and explanatory variables across the 672 clusters of the 2017/18 BDHS. Survey weights were applied using svy command in Stata to calculate the proportions. The estimated proportions were then merged with the spatial locations of the BDHS clusters. The spatial autocorrelation (Global Moran’s I) statistic was used to assess whether the pattern of non-use of modern contraception was dispersed, clustered or randomly distributed in the study area. The Gettis-ord Gi^*^ was calculated to measure how the spatial autocorrelation differed through the study locations by computed Gi statistics for each area. Z-score and the relevant p-values were then calculated to determine the statistical significance in the clustering of non-use of modern contraception, including hot spot (area where modern contraception use was low) and cold spot (area where modern contraception use was high). We considered a False Discovery Rate (FDR) correction while using the Getis-Ord G^*^i (d) statistics to account for multiple dependent tests. The importance of considering the FDR correction method in DHS data has been described elsewhere [23]. The geographically weighted regression (GWR) model was used to determine the cluster wise coefficients of the explanatory variables of the non-use of modern contraception. The variables that fitted the assumptions of GWR were included in the model and selected using exploratory regression and ordinary least square regression (OLS) models. We first ran the exploratory regression model to identify variables to be included in the OLS model while the OLS model was used to identify the predictors of the observed spatial pattern of non-use of modern contraception in Bangladesh. Through the OLS model an overall coefficient of the explanatory variables of non-use of modern contraception was produced while the GWR produced cluster wise coefficients. We performed all descriptive analysis using the Stata software version 15.1 (Stata corporation, college station, Texas, USA). The statistical package R (version 4.1.1) was used for all other analyses (i.e. exploratory regression, OLS and GWR). All methods were performed in accordance with the relevant guildelines and regulations.

### Ethical consideration

We analysed secondary data extracted from the Demography and Health Survey (DHS) program in de-identified form with permission to analyse. The survey was approved by the National Research Ethics Committee of Bangladesh and ICF Macro International. No other ethical approval is required to analyse this survey data. Informed written consent was obtained from all participants.

## Results

### Socio-demographic and community characteristics of the respondents

Of the 20,127 women interviewed, 16,135 met the inclusion criteria, and were included in the analysis (80% of the main survey participants). The median (IQR) age of the women was 25 (21-29) years with almost half having three or more children (Table 1). About one-fifth of the participants (19.2%) had received no formal education and half were currently employed. Around 72% of the women reported that they lived in a rural area while about 25%, 17% and 14% of women resided in Dhaka, Chattogram and Rajshahi divisions, respectively.

**Table 1:**
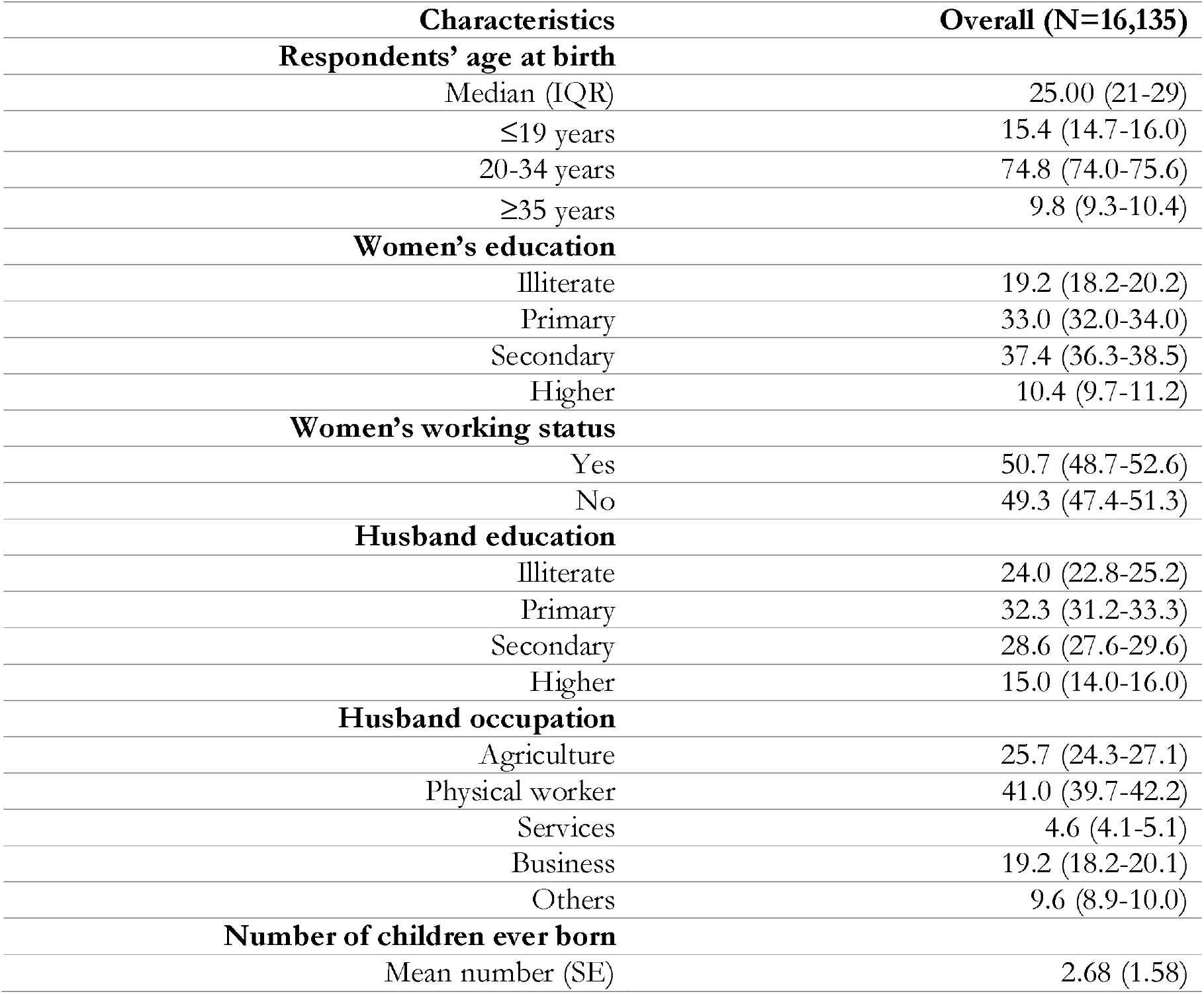
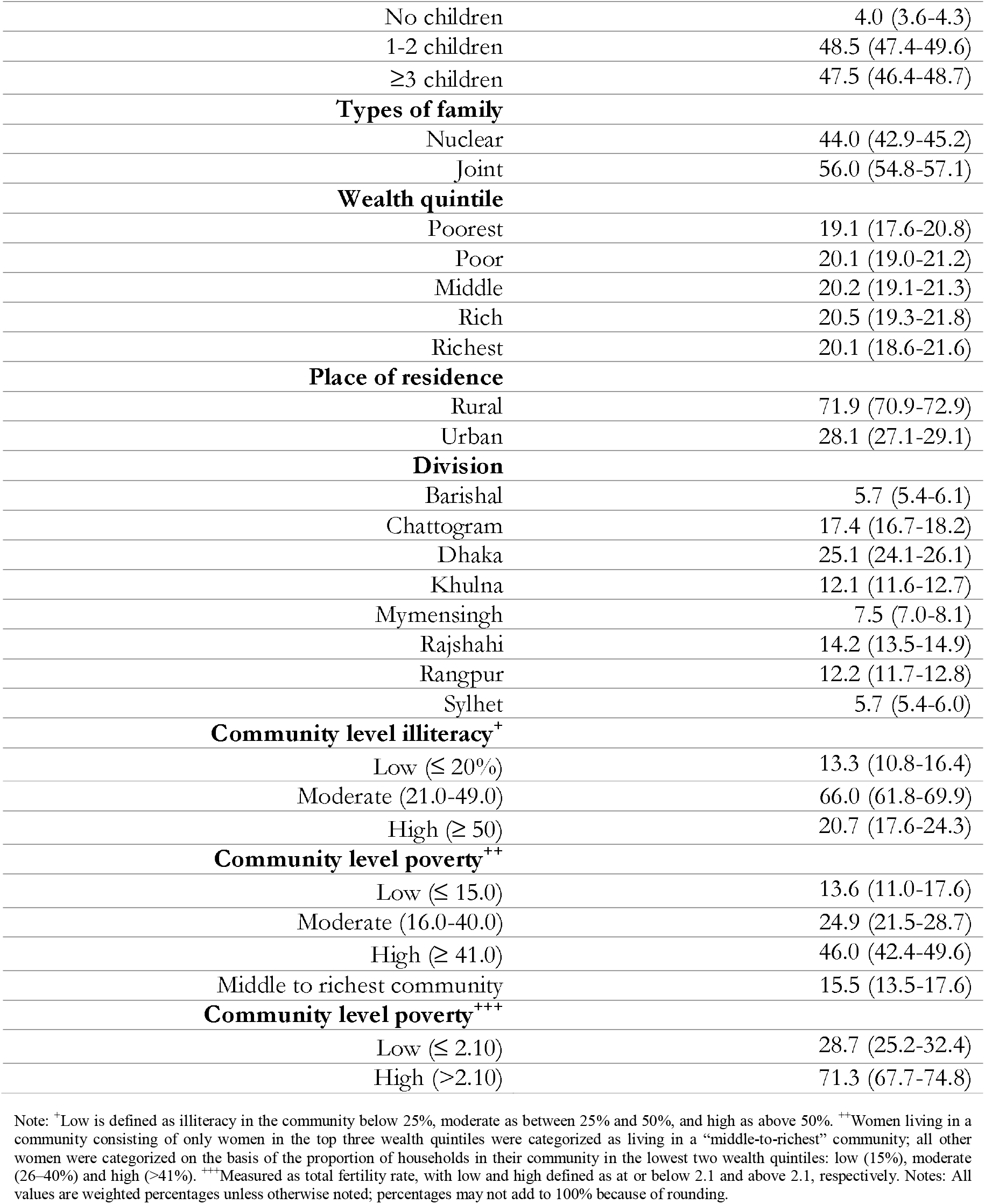
Characteristics of respondents, Bangladesh 2017/18.

| Characteristics | Overall (N=16,135) |
| --- | --- |
| **Respondents' age at birth** |  |
| Median (IQR) | 25.00 (21-29) |
| ≤19 years | 15.4 (14.7-16.0) |
| 20-34 years | 74.8 (74.0-75.6) |
| ≥35 years | 9.8 (9.3-10.4) |
| **Women's education** |  |
| Illiterate | 19.2 (18.2-20.2) |
| Primary | 33.0 (32.0-34.0) |
| Secondary | 37.4 (36.3-38.5) |
| Higher | 10.4 (9.7-11.2) |
| **Women's working status** |  |
| Yes | 50.7 (48.7-52.6) |
| No | 49.3 (47.4-51.3) |
| **Husband education** |  |
| Illiterate | 24.0 (22.8-25.2) |
| Primary | 32.3 (31.2-33.3) |
| Secondary | 28.6 (27.6-29.6) |
| Higher | 15.0 (14.0-16.0) |
| **Husband occupation** |  |
| Agriculture | 25.7 (24.3-27.1) |
| Physical worker | 41.0 (39.7-42.2) |
| Services | 4.6 (4.1-5.1) |
| Business | 19.2 (18.2-20.1) |
| Others | 9.6 (8.9-10.0) |
| **Number of children ever born** |  |
| Mean number (SE) | 2.68 (1.58) |
| No children | 4.0 (3.6-4.3) |
| 1-2 children | 48.5 (47.4-49.6) |
| ≥3 children | 47.5 (46.4-48.7) |
| **Types of family** |  |
| Nuclear | 44.0 (42.9-45.2) |
| Joint | 56.0 (54.8-57.1) |
| **Wealth quintile** |  |
| Poorest | 19.1 (17.6-20.8) |
| Poor | 20.1 (19.0-21.2) |
| Middle | 20.2 (19.1-21.3) |
| Rich | 20.5 (19.3-21.8) |
| Richest | 20.1 (18.6-21.6) |
| **Place of residence** |  |
| Rural | 71.9 (70.9-72.9) |
| Urban | 28.1 (27.1-29.1) |
| **Division** |  |
| Barishal | 5.7 (5.4-6.1) |
| Chattogram | 17.4 (16.7-18.2) |
| Dhaka | 25.1 (24.1-26.1) |
| Khulna | 12.1 (11.6-12.7) |
| Mymensingh | 7.5 (7.0-8.1) |
| Rajshahi | 14.2 (13.5-14.9) |
| Rangpur | 12.2 (11.7-12.8) |
| Sylhet | 5.7 (5.4-6.0) |
| **Community level illiteracy<sup>+</sup>** |  |
| Low (≤ 20%) | 13.3 (10.8-16.4) |
| Moderate (21.0-49.0) | 66.0 (61.8-69.9) |
| High (≥ 50) | 20.7 (17.6-24.3) |
| **Community level poverty<sup>++</sup>** |  |
| Low (≤ 15.0) | 13.6 (11.0-17.6) |
| Moderate (16.0-40.0) | 24.9 (21.5-28.7) |
| High (≥ 41.0) | 46.0 (42.4-49.6) |
| Middle to richest community | 15.5 (13.5-17.6) |
| **Community level poverty<sup>+++</sup>** |  |
| Low (≤ 2.10) | 28.7 (25.2-32.4) |
| High (>2.10) | 71.3 (67.7-74.8) |

Note: <sup>+</sup>Low is defined as illiteracy in the community below 25%, moderate as between 25% and 50%, and high as above 50%. <sup>++</sup>Women living in a community consisting of only women in the top three wealth quintiles were categorized as living in a “middle-to-richest” community; all other women were categorized on the basis of the proportion of households in their community in the lowest two wealth quintiles: low (15%), moderate (26–40%) and high (>41%). <sup>+++</sup>Measured as total fertility rate, with low and high defined as at or below 2.1 and above 2.1, respectively. Notes: All values are weighted percentages unless otherwise noted; percentages may not add to 100% because of rounding.

### Geographical variation of non-use of modern contraception in Bangladesh

The geographical distribution of modern contraception non-use in Bangladesh is presented in Table 2. In all, 44% (42.5–45.3) and 39% (37.7–41.3) of the women living in rural and urban areas reported non-use of modern contraception, respectively. Among the eight administrative divisions, the highest proportion of women living in the Sylhet division (50.8%, 47.7-53.8) reported modern contraception non-use, followed by women living in the Chattogram division (49.9%, 46.9-53.0).

**Table 2:** Weighted proportion of non-use of modern contraception in Bangladesh by place of residence and administrative division, BDHS 2017/18 (N=16,135)

|  | Non-use of modern contraception n (%<br>95%CI) | P-value |
| --- | --- | --- |
| Overall | 6, 886 (42.8, 41.6-43.8) |  |
| Place of residence |  |  |
| Urban | 1791 (39.5, 37.7-41.3) | *p<0.001* |
| Rural | 5095 (43.9, 42.5-45.3) |  |
| Administrative Division |  |  |
| Barishal | 424 (45.8, 42.4-49.2) | *p<0.001* |
| Chattogram | 1405 (49.9, 46.9-53.0) |  |
| Dhaka | 1662 (41.0, 38.3-43.8) |  |
| Khulna | 856 (43.8, 41.3-46.4) |  |
| Mymensingh | 453 (37.3, 34.7-40.1) |  |
| Rajshahi | 907 (39.7, 36.7-42.7) |  |
| Rangpur | 714 (36.2, 33.2-39.2) |  |
| Sylhet | 465 (50.8, 47.7-53.8) |  |

### Spatial variation in non-use of modern contraception

A significant positive autocorrelation with the Global Moran’s I index of 0.379 (*p-value <0*.*001*) was found between non-use of modern contraception and geographic location. The Getis-Ord General G statistics revealed that the presence of high clustering (z-score= 5.78, *p<0*.*001*). The hot spots of non-use of modern contraception were mostly located in the Sylhet, Chattogram and part of the Barishal divisions. The cold spot of non-use of modern contraception were mostly located in the Rangpur, Mymensingh, Rajshahi and Dhaka divisions (Figure 1).

**Figure 1:**
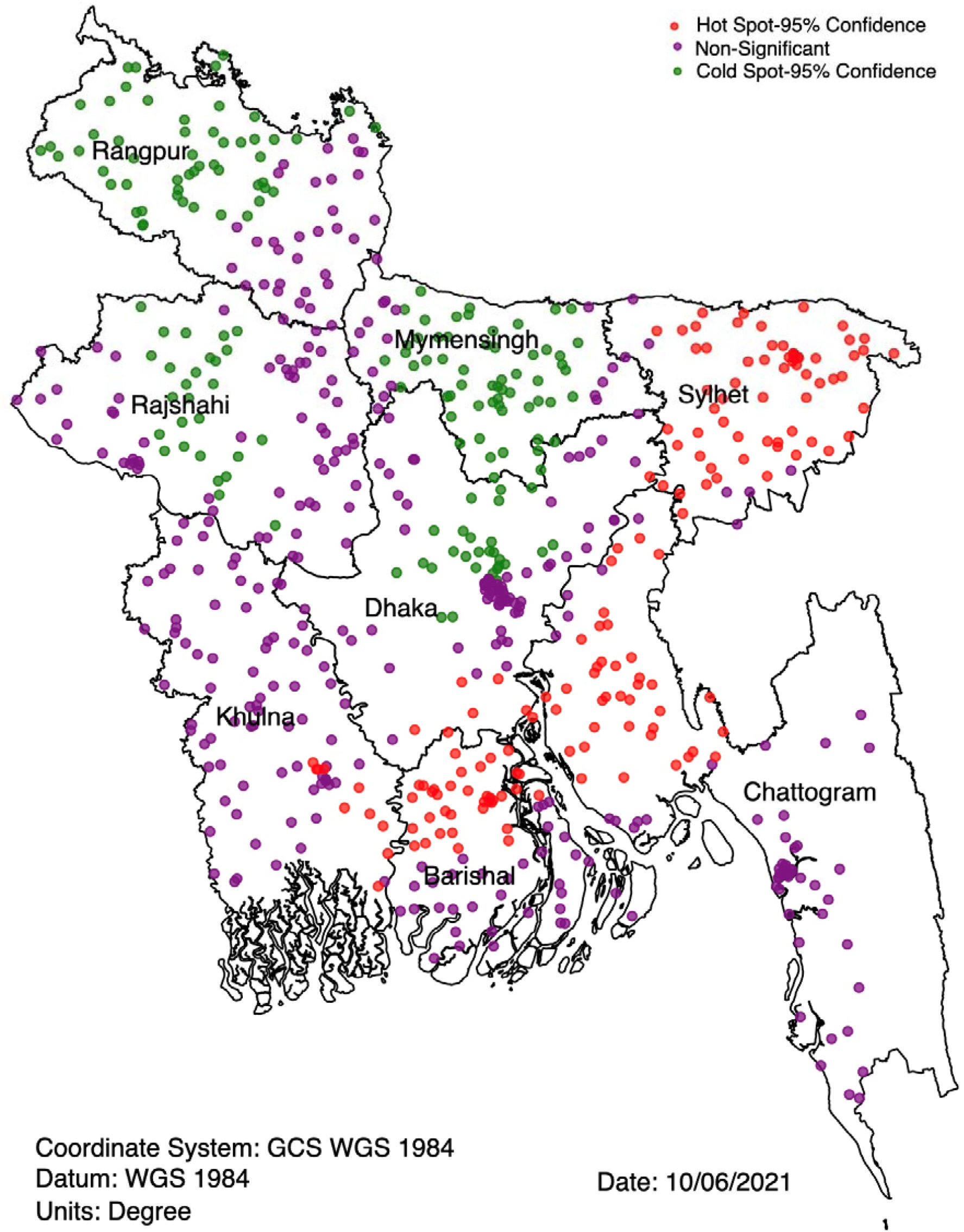
Hot spots and cold spots of non-use of modern contraception in Bangladesh, 2017/18 BDHS

### Predictors of non-use of modern contraception

All individual, household and community level factors considered were first added in the exploratory regression analysis to get a precise list of factors to be included in the OLS. There was no evidence of multicollinearity in the included variables. From this modelling, five variables, of which there were eleven components, had a positive relationship with the non-use of modern contraception (see Table 3). These variables were then included in the GWR. The summary of this model is presented in Table 4, and the cluster wise coeffeicients are plotted graphically for the individual (Figure 2[a–c]), household-level (Figure 3[a–f]) and community level factors (Figure 4[a–b]).

**Table 3:** Ordinary least square regression model identifying significant factors of short birth interval in Bangladesh, BDHS 2017/18

| Variable category | Coefficient | Standard error | t-statistics | Probability | Robust std-error | Robust t-statistics | Robust probability | VIF |
| --- | --- | --- | --- | --- | --- | --- | --- | --- |
| Illiterate women | 0.06 | 0.11 | 53.0 | <0.01 | 54.0 | 53.4 | <0.01 | 5.64 |
| Primary educated women | 0.23 | 0.10 | 12.0 | <0.01 | 13.0 | 12.8 | <0.01 | 6.16 |
| Secondary educated women | 0.10 | 0.93 | 37.0 | <0.01 | 40.0 | 36.9 | <0.01 | 5.42 |
| Women's husband were illiterate | 0.20 | 0.10 | 19.7 | <0.01 | 20.0 | 20.2 | <0.01 | 6.92 |
| Women's husband were primary educated | 0.14 | 0.09 | 16.0 | <0.01 | 16.8 | 16.4 | <0.01 | 4.97 |
| Women's husband were secondary educated | 0.08 | 0.09 | 80.0 | <0.01 | 83.0 | 79.3 | <0.01 | 4.14 |
| Women's husband were physical worker | 0.06 | 0.04 | 15.7 | <0.01 | 15.7 | 16.0 | <0.01 | 1.29 |
| Women's husband were services holder | 0.08 | 0.11 | 71.0 | <0.01 | 68.0 | 69.9 | <0.01 | 3.68 |
| Women's husband were businessmen | 0.11 | 0.05 | 22.6 | <0.01 | 22.3 | 23.0 | <0.01 | 1.26 |
| Moderate community level illiteracy | 0.12 | 0.01 | 18.0 | <0.01 | 17.6 | 18.1 | <0.01 | 1.47 |
| Moderate to richest community | 0.07 | 0.02 | 46.9 | <0.01 | 51.1 | 46.4 | <0.01 | 1.10 |
| **Model diagnostics** |  |  |  |  |  |  |  |  |
| Number of observation (EAs) | 672 | Akaike's Information Criterion (AIC): |  |  |  |  | 1018.13 |  |
| Multiple R-square | 0.83 | Adjusted R-square |  |  |  |  | 0.82 |  |
| Joint F-Statistics | 1692.13 | Probability (> F), (11,669) degrees: |  |  |  |  | <0.01 |  |
| Joint Wald Statistics | 158.23 | Probability (> chi-squared) |  |  |  |  | <0.01 |  |
| Koenker (BP) Statistics | 1862.14 | Probability (> chi-squared) |  |  |  |  | <0.01 |  |
| Jarque-Bera Statistics | 0.86 | Probability (> chi-squared) |  |  |  |  | <0.01 |  |

**Table 4:**
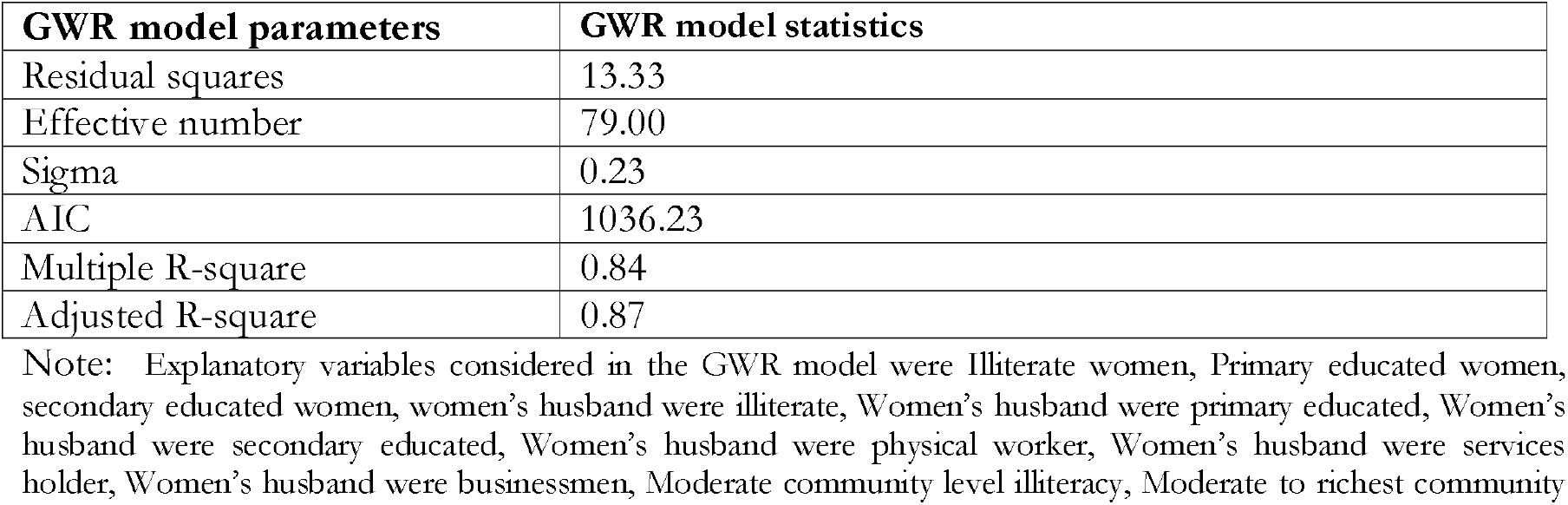
Geographically weighted regression model assessing factors of non-use of modern contraception in Bangladesh, BDHS 2017/18.

| **GWR model parameters** | **GWR model statistics** |
| --- | --- |
| Residual squares | 13.33 |
| Effective number | 79.00 |
| Sigma | 0.23 |
| AIC | 1036.23 |
| Multiple R-square | 0.84 |
| Adjusted R-square | 0.87 |

Note: Explanatory variables considered in the GWR model were Illiterate women, Primary educated women, secondary educated women, women's husband were illiterate, Women's husband were primary educated, Women's husband were secondary educated, Women's husband were physical worker, Women's husband were services holder, Women's husband were businessmen, Moderate community level illiteracy, Moderate to richest community

**Figure 2.**
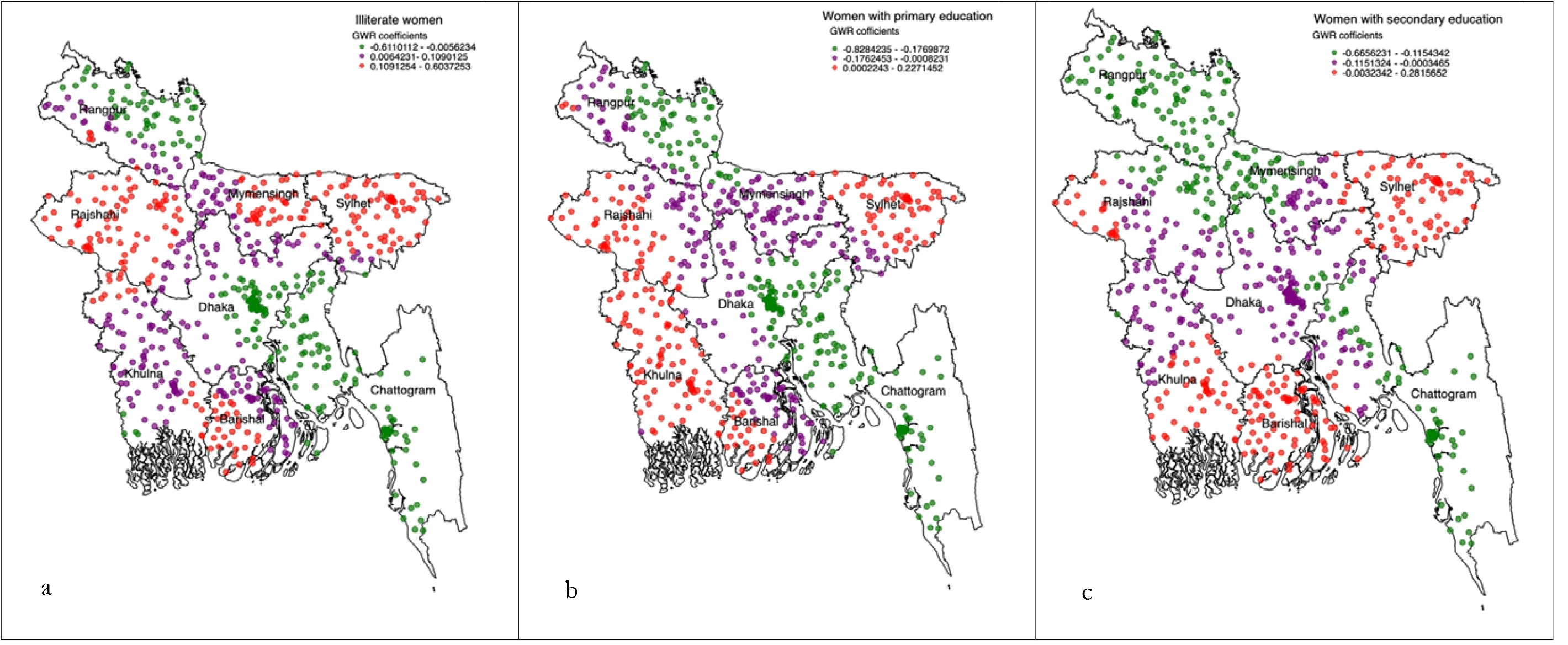
[a–c]: Individual level predictors of non-use of modern contraception accessed through using the geographically weighted regression model

**Figure 3.**
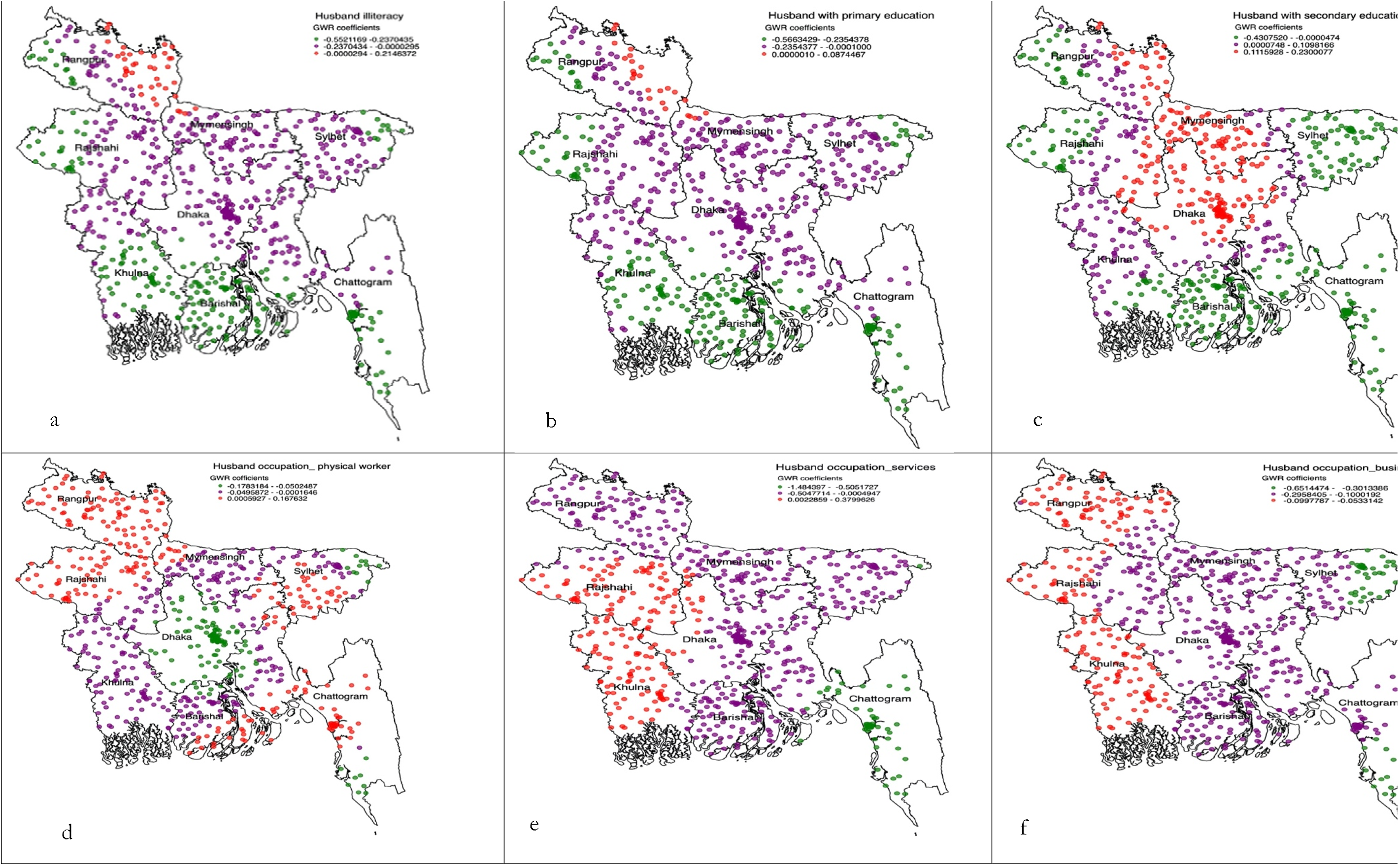
[a–f]: Household level predictors of non-use of modern contraception accessed through using geographical weighted regression model

**Figure 4.**
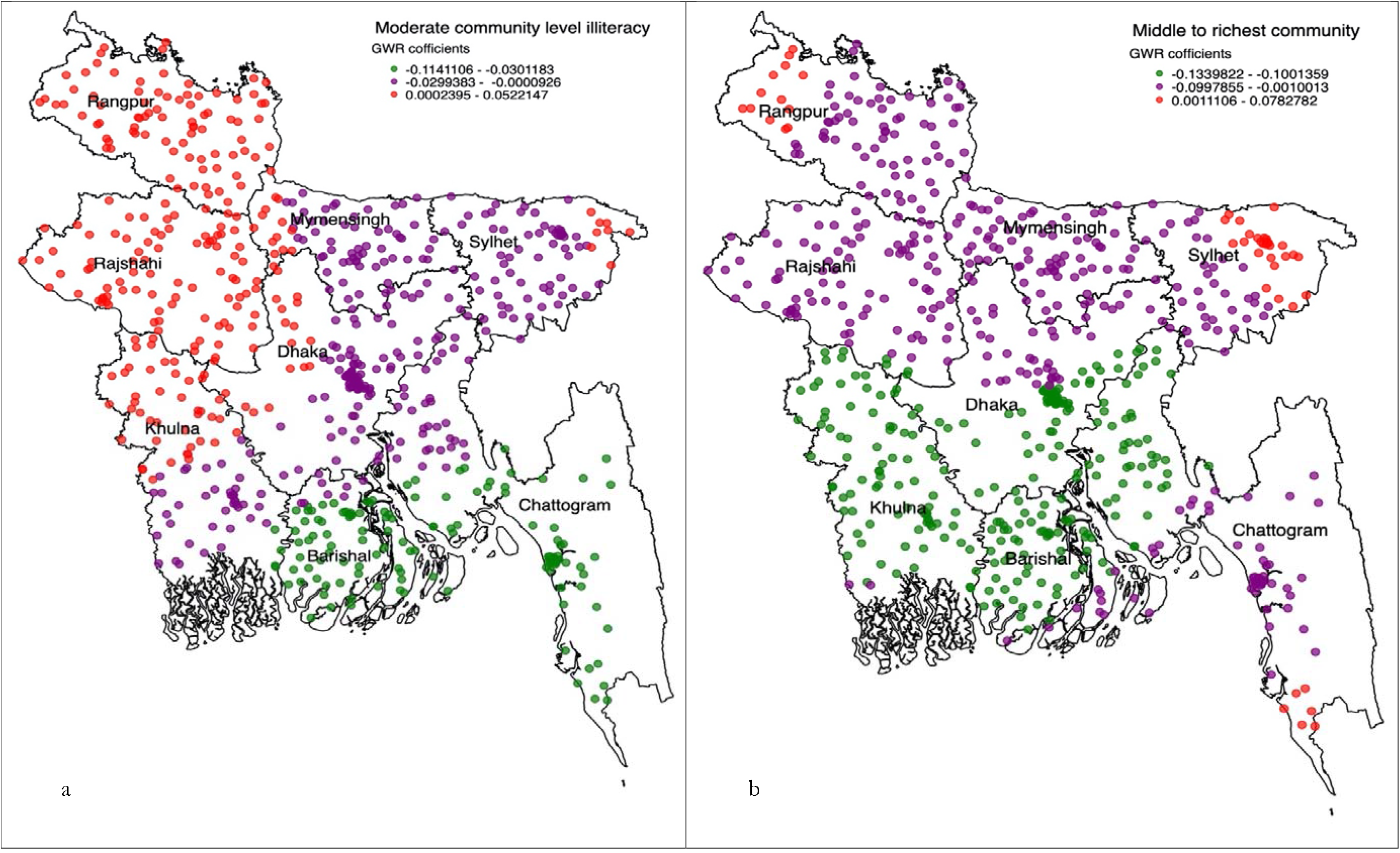
[a–b]: Community level predictors of non-use of modern contraception accessed through using the Geographical weighted regression model

In the Sylhet division, where the majority of non-use of modern contraception hot spots were located, significant predictors of non-use of modern contraception were women’s education up to the secondary level (Figure 2 [a–c]). In the other two hot spot prevalent divisions, Barishal and Chattogram, non-use of modern contraception was found to be lower among primary and secondary educated women. Women whose husbands work involved physical labour were more likely reported non-use of modern contraception, a relationship which was true for all hot spot prevalent areas in the Sylhet, Barishal, and Chattogram divisions. The lower likelihood of non-use of modern contraception was found among women whose husbands were secondary level educated and resided in moderate community level literacy and moderate to richest community (4 [a–b]).

In the cold spots prevalent divisions, Mymensingh and Rajshahi, the likelihood of modern contraception non-use was found lower among women whose husbands were either illiterate or primary educated (3 [a-b]). Women whose husbands were secondary educated reported a higher likelihood of non-use of modern contracetion in all cold spots prevalent areas: Mymensingh, Rajshahi, and Rangpur (3 [c]). Moreover, in Rajshahi and Rangpur divisions, the non-use of modern contraception was found to be higher among women whose husbands were physical workers and engaged with business. However, in the Khulna division, higher likelihoods of non-use of modern contraception were reported among women whose husbands were enaged with either services or business. Conversely, this association was found to be the opposite for cold spots areas of the Mymensingh division, where lower likelihoods of non-use of modern contraception were reported among women whose husbands’ work involved either physical labour or business or service. We also found a lower likelihood of non-use of modern contraception among women who resided in the community with moderate illiteracy and those who had middle to richest poverty (4[a–b]).

In the Dhaka and Khulna divisions, illiterate women reported a higher likelihood of non-use of modern contraception, whereas primary educated and secondary educated women reported lower likelihoods of non-use of modern contraception. Higher community level illiteracy was found to be associated with increase likelihoods of non-use of modern contraception in Dhaka, Khulna, Rangpur, and Rajshahi divisions. Lower likilihoods of non-use of modern contraception were found lower among women resided in the moderate to richest community-level poverty.

## Discussion

This is the first study conducted in a LMICs and in particular Bangladesh, to explore the geographical variation using sophisticated analysis technique to identify areas where modern contraception non-use was clustered. Individual, household, and community-level factors associated with the higher and lower use of modern contraception were also determined for each cluster. We found that almost all clusters where modern contraception use was low were located in the Sylhet, Barishal and Chattogram divisions. Factors associated with the non-use of modern contraception were found to vary across clusters, except for the individual level factors women’s literacy and husband’s education up to secondary level. These factors were found to be predictive of non-use of modern contraception in general in Bangladesh. The findings suggest a clear need for area-specific policies and programs to increase modern contraception use in Bangladesh.

Importantly, our study found that around 43% of the women in Bangladesh do not use modern contraception [24]. This percentage is lower than the average prevalence of modern contraception use in LMICs [25], however, comparable with the available studies in Bangladesh [26]. We also found the divisional variations of modern contraception use, with modern contraception was found to be lower in the Sylhet, Chattogram and Barishal divisions and higher in the Rangpur, Mymensingh and part of the Rajshahi division. Previous studies in Bangladesh also reported a higher odd of modern contraception non-use in the Sylhet, Barishal and Chattogram divisions and lower odds of non-modern contraception in the Rangpur, Mymensingh and Rajshahi divisions [18, 27]. However, our findings were based on cluster level (smallest area under division) estimates which enable us to identify specific areas of contraception unmet need.

We found women’s illiteracy and husband’s education up to secondary level to be associated with modern contraception non-use in each of the areas, regardless of their clustering status. This indicates, that although education plays an important role in modern contraception use or non-use in Bangladesh, clustering of non-use of modern contraception may be attributed to other factors where education also plays an important role. While further research is required to tease out these important relationships, these findings point to a need for revising the current policies and programs to create awareness among women and their husbands regarding the importance of using modern contraception. The government of Bangladesh employs two approaches to the provision of contraception education to eligible couples: (i) dissemination of information through mass media, and (ii) individual contraceptive counselling through family planning visits [28, 29]. In the recent five-year plan, the inclusion of women’s husbands in family planning counselling is specified, however, at the field level, this has mostly not been implemented [30]. These ways of disseminating modern contraceptive use information bring some major challanges, particularly among uneducated or less educated populations. For instance, information disseminated through the mass media (such as radio and television) is one way, therefore, illiterate or less educated people mostly do not benefit from it [31]. This burden is even higher for working women as there is the possibility of a mismatch between the information disseminated and women’s availability to join these programs [17]. Women may also not be able to buy radios or televisions [32, 33]. While individual contraception counselling taiolered to the needs of women may play an effective role in overcoming these challenges, this type of contraceptive counselling has been declining of the past 20 years [5]. As such, field level monitoring of such programs would be required.

We found that women whose husbands work in physical labour are less likely to use modern contraception, whereas women whose husbands are businessmen and services holders reported a mixed finding across hwith higher or lower use of modern contraception. In the case of physical labour, the observed association is easily understandable as they are mostly illiterate as well as agricultural workers among which the knowledge over the importance of using modern contraception is low [34]. Moreover, people in this group in Bangladesh tend to have more children because of the potential increase of income and assistance [35]. They are also influenced by several religious and community level misconceptions. For instance, Muslims in Bangladesh often believe their religion Islam, the religion of more than 90% of the total population in Bangladesh, do not support contraception use and controlling fertility through contraception is equivalent to the killing of a human being [36]. These sorts of misconceptions are even more common among illiterate, and people engaged in marginalized jobs. In addition, at the community level, there is a misconception that the use of contraception is risky for the health of the couple, particularly women, and often an important risk factor of infertility [17]. These social taboos and misconceptions may be the reasons for getting mixed findings of contraception use among women whose husbands are businessmen or service holders, as depending on the communities and areas where women resided, the severity of these taboos varies [17].

Community-level illiteracy is found as an important predictor of non-use of modern contraception, mostly in the Rajshahi, Rangpur and Khulna divisions—the areas where modern contraception use was found to be high. However, surprisingly in the Sylhet, Barishal and Chattogram divisions, where prevalence modern contraception non-use was found to be high, community-level illiteracy was found to facilitate non-use. This finding is aligned with the area level education as per the most recent available estimate in Bangladesh [37]. However, there might be some other factors influencing the relationship. For instance, reproductive health indicators have constantly been reported poor in the Sylhet, Barishal and Chattogram divisions [5]. Therefore, governmental and non-governmental organizations have implemented several programs in these divisions, mainly in the era of Millennium Development Goals (2000-2015) as well as the SDGs, where the major target group were disadvantaged communities [38]. These may bring this change; however, we recommend a further study to explore the reasons behind this reported association. The association between the community level poverty and non-use of modern contraception reported in this study is straightforward and could be easily explained as people in Bangladesh often live in a cluster with other people in similar backgrounds. Therefore, people in the middle to the richest community are often educated and engaged in a better job, which make them self-aware to use modern contraception.

This study has several strengths and limitations. As far as we know, this is the first study in Bangladesh as well as LMICs that explored areas of higher and lower use of modern contraception. The explanatory variables were selected carefully through a comprehensive review of the previous papers and included in the model through proper model building techniques. Therefore, the findings of this study will have practical implications to develop area level policies and programs. However, the major limitation of this study is the analysis of data from a cross-sectional survey, therefore the findings are correlational only not causal. In addition, to protect the privacy of the respondents, the BDHS relocated the cluster location in the map up to 5 km for the urban sample and 2 km for the rural sample. A further 2% of the total sample cluster location displaced up to 10 km. Therefore, the cluster location shown on the map and their risk factors are different from the actual location. However, during displacing the BDHS ensured displaced locations are placed in the same administrative boundary. Therefore, the findings reported in this study are still valid and should be used to develop regional-level policies and programs.

## Conclusion

This study reported that 43% of women in Bangladesh do not use modern contraception with a significant variation identified across divisions. Areas where modern contraception use was found to be low were mostly located in the Sylhet, Barishal and part of Chattogram divisions. Modern contraception use was found to be higher in the Rangpur, Rajshahi, and Mymensingh divisions. The risk factors of non-use of modern contraction varied throughout the country, except for lower education of women and their husbands. Community-level illiteracy played different roles depending upon the area. The findings therefore support the need for the design and implementation of area-specific policies and programs rather than the generalised policies and programs to increase modern contraception use currently being administered in Bangladesh.

## Data Availability

All data produced are available online at

https://dhsprogram.com/data/

## Acknowledgement

The authors would like to thank the MEASURE DHS for granting access to the 2011 and 2017/18 BDHS data.

## Authors’ contributions

MNK designed the study, performed the data analysis, and wrote the first draft of this manuscript. MNK and MLH critically reviewed and edited the previous versions of this manuscript. All authors approved this final version of the manuscript.

## Data availability

We analysed third party data available in the Measure DHS website: https://dhsprogram.com/data/available-datasets.cfm. <u>Anone interested can download the dataset from this website by submitting a research proposal. We could share the data and make it available in the data respiratory.</u>

